# COVID-19 Network Model to Evaluate Vaccine Strategies towards Herd Immunity

**DOI:** 10.1101/2020.12.22.20248693

**Authors:** Josephine N. A. Tetteh, Van Kinh Nguyen, Esteban A. Hernandez-Vargas

## Abstract

With more than 60 million confirmed cases and more than 1.5 million deaths, SARS-CoV-2 has paralyzed our societies, leading to self isolation and quarantine for several months. A COVID-19 vaccine remains a critical element in the eventual solution to this public health crisis. Two vaccines are ready to be mass produced and eventually supplied to the population. Here, we develop an epidemiological network model able to represent COVID-19 pandemic dynamics of different countries such as in Italy. Stochastic computational simulations identify the necessary number of vaccines and vaccine efficacy thresholds capable of preventing an epidemic whilst adhering to lockdown guidelines. Assuming a vaccine efficacy of 100% in a mass vaccination program, at least 60% of a given population should be vaccinated to obtain herd immunity. Otherwise, new waves of COVID-19 would be reported. However, reaching 60% of the population will be an unprecedented mass immunisation campaign that will take several months and millions of dollars. If the vaccine efficacy reports lower levels of efficacy in practice, then the coverage of vaccination would be needed to be even higher. Simulations suggest that the “Ring of Vaccination” strategy, vaccinating susceptible contact and contact of contacts, would be a more tractable vaccine strategy to prevent the new waves of COVID-19 meanwhile a high percent of the population is vaccinated.

## 1 Introduction

The novel Coronavirus SARS-CoV-2 epidemic which first emerged from Wuhan, China in December 2019 has spread globally, causing high levels of mortality and morbidity worldwide. To curb the spread, governments across the world have implemented measures ranging from quarantining, social distancing, wearing of face masks among others. Amidst this crisis, national health care systems such as in Italy and the United States of America have been overwhelmed by the ever-increasing number of infection cases [36].

COVID-19 epidemiological models have been formulated to understand and curb the spread of the disease. Many of these models follow an SIR framework either in the deterministic or stochastic form or both [2, 59, 53, 55, 9, 10]. Other variations and modifications to this general model have been considered including SEIR [47, 30, 50] and SIRD [13] compartmental models. Some models also include parameters such as age-heterogeneity [56], guiding the flow of users in supermarkets [19], and governmental policies [50, 4, 32]. In addition, a few studies incorporate the dynamics of the disease within an individual host [21, 1]. However, only a few of these models, currently predicting the SARS-CoV-2 pandemic, consider the structure of the population and the underlying interactions between individuals [15, 61, 22].

The assumption of random homogeneous mixing in epidemiological models has been doc-umented to be unrealistic in nature as populations have underlying structural properties and individuals tend to interact with each other [14]. Increasingly, network theory is being used in epidemiology [35, 26, 51, 16]. In particular, social networks have gained popularity in conceptualising the effects of social interaction during epidemics in a given population [38, 25, 54]. Contacts between individuals in a given population can be captured in a network where nodes represent individuals and the edges represent the connections between them [20]. Social networks are thus important determinants of infectious disease transmission as for example, infections transmitted by close contact can easily spread along the paths of a network.

With the continuation of SARS-CoV-2 in many parts of the world, the push for a vaccine has become highly necessary. Pharmaceutical companies are in a race to develop suitable vaccines as there is a lack of other alternatives. As of October 2020, there were 17 candidate vaccines undergoing trial at various stages. Owing to the fact that it is a novel viral disease, it is still unclear what levels of vaccine efficacies will be sufficient to curb the spread of the virus. Identifying such efficacies earlier can direct vaccine development and administration in the population [23]. Previous studies suggest that vaccination would be effective for protecting the host against SARS-CoV-2 [28], however, there have not been studies to evaluate the potential effects of different vaccination programs over a network model.

A key question to be answered is, how much vaccine is required to create herd immunity to block SARS-CoV-2 transmission? [3]. In other words, how many people need to be vaccinated in order to reach herd immunity? Here, we employ a network-based approach to explore the potentials of two vaccination schemes, classical mass vaccination and ring vaccination, in minimizing the spread of SARS-CoV-2, see Figure 1. Ring vaccination is a vaccination strategy in which infected cases and contacts of cases are identified and vaccinated [18, 39]. Our analysis uses a stochastic network simulation model of SARS-CoV-2 transmission to examine its control by mass and ring vaccination strategies with varying efficacies in the presence of non-pharmaceutical disease control interventions. Given that vaccinating millions of people will require a lot of time, this study implements a lockdown period as a further control measure during vaccination strategies in our simulations. The main objective is to identify the necessary vaccine efficacy thresholds capable of preventing an epidemic whilst adhering to lockdown instructions.

**Figure 1:**
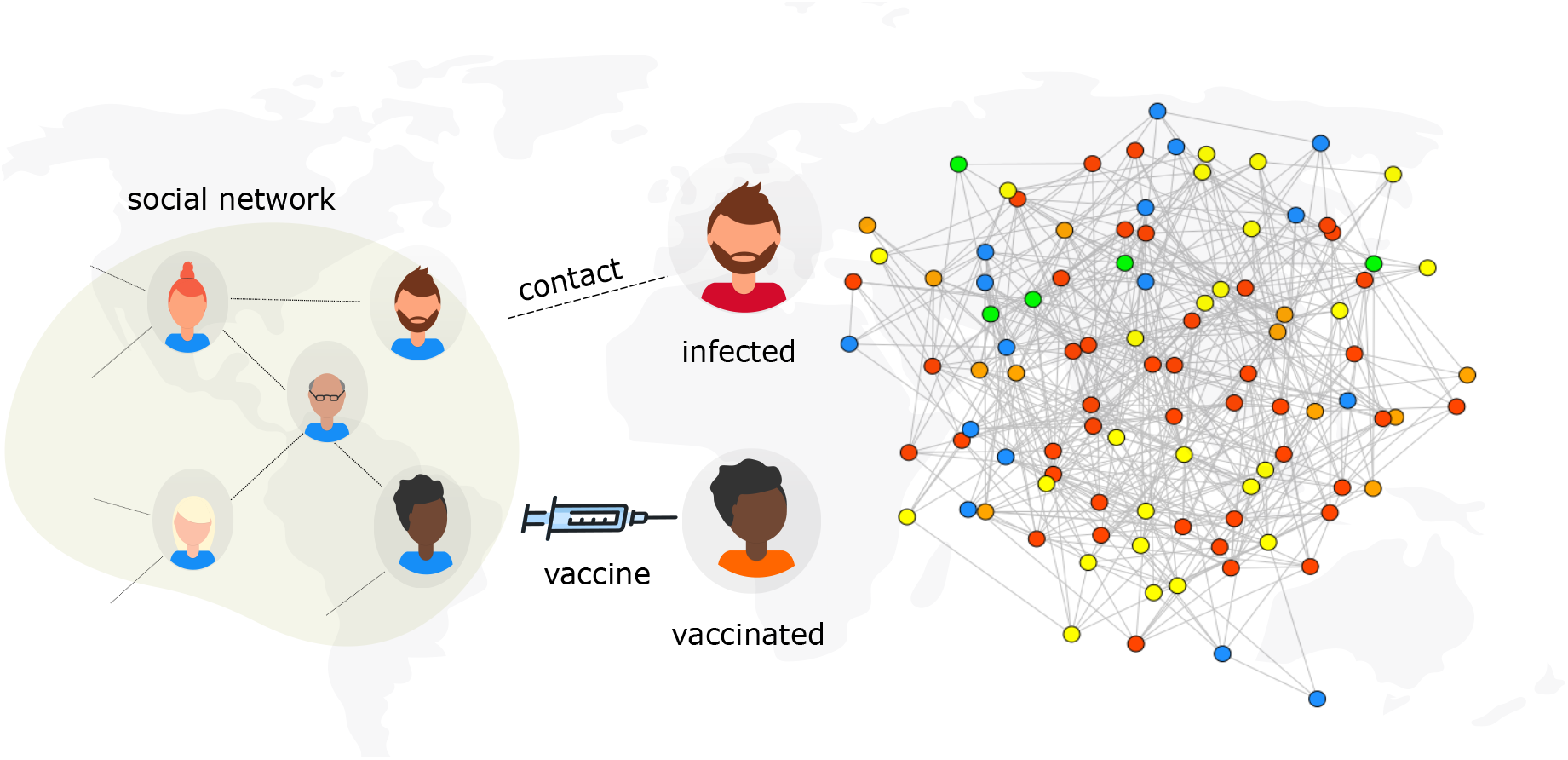
Model scheme. An illustration of infection spread with vaccination on a social network. Individuals in the social network are considered as nodes and contact between nodes exposed to the virus and those who do not have the virus can potentially lead to a transmission. Persons who are vaccinated are considered to be immune to infection. The epidemic state of each node is represented by a colour: Susceptibles, *S*, (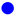), Exposed, *E*, (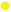), Infected, *I*, (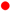), Vaccinated, *V*, (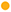) and Recovered, *R*, (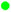).

## 2 Modelling Epidemic Spread on Networks

### 2.1 Model Setup

#### Network Generation

To study the impact of vaccination on the epidemic dynamics of SARS-CoV-2, the underlying structure of human interactions is represented by a typical complex network. As heterogeneous networks are often used to explore epidemic spread, we consider the infection dynamics on an Erdos-Renyi (ER) network due to its tractability and practicability [27, Chapter 1]. The ER network is based on the *G*(*n, M*) random graph model characterized by two parameters; the network size *n* and the number of edges *M* which assigns exactly *M* number of edges to each graph. In the ER networks used for this study, *n* = *N, M* = 5*N*, that is *G*(*N*, 5*N*), which yields an average degree distribution ⟨ *k*⟩ = 10.

#### Epidemic Spread

SARS-CoV-2 is a disease which spreads primarily through close contact with an infected person. Following the exposure to the SARS-CoV-2 virus and before symptom onset, individuals go through an incubation period of about 14 days with the average being about 5 − 6 days [33, 52, 7]. After this incubation period, infected individuals become symptomatic and are able to transmit to others through respiratory droplets or by direct contact [33].

#### Epidemic States

We simulated a Susceptible - Exposed - Infected - Recovered - Vaccinated (SEIRV) process on an Erdos-Renyi network with *N* = 10^6^ nodes. At any instant during the infection process, the status of a node can be in any one of five possible states: Susceptible (*S*, not infected but can be infected), Exposed (*E*, infectious, may not show symptoms and can transmit to others), Infected (*I*, infectious and symptomatic, capable of transmission to others), Recovered (*R*, recovered and immune to the disease) and Vaccinated (*V*, vaccinated). Therefore, at any time, *t*, in the infection process, *N* (*t*) = *S*(*t*) + *E*(*t*) + *I*(*t*) + *R*(*t*) + *V* (*t*).

#### Epidemic Process

The model (depicted in Algorithm 1 proceeds in discrete one-day time steps for a period of 360 days to determine disease dynamics. Simulation codes can be found in Supplemental Materials. At the initial state of the epidemic process, all individuals in the network are susceptible except one (patient zero) which is in the exposed state. On each day during the epidemic process, there is interaction between individuals and infected persons can potentially transmit to their susceptible contacts. If a susceptible individual comes into contact with someone who has the virus (that is, a person in the *E* or *I* state), a Binomial trial is used to determine if the contact results in an infection. If yes, the newly infected susceptible individual moves from the *S* state to the *E* state. Any individual exposed to the disease remains in the *E* state for the duration of the incubation period after which it moves to the *I* state. Infectious individuals can transmit to their neighbours, when they come into contact, in Binomial trial with a given probability, *β*(*t*) and then move into the *R* state after the infectious period.

In the course of an epidemic, the rate of infection is never constant. As interventions are being executed, the rate of infection decreases, otherwise it increases. To model this response, we utilise a double logistic function to model the various phases of the infection dynamics leading to a decline in cases when interventions are initiated and a resurgence in cases when interventions cease. The double logistic function used to define a time dependent probability of infection, *β*(*t*) is as follows:

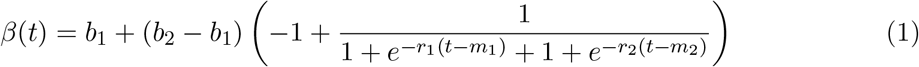

where *b*_1_ is the first boundary (i.e. function value at time zero), *b*_2_ is the second boundary, *r*_1_ is the rate of change of first period, *r*_2_ is rate of change of second period, *m*_1_ is the midpoint of the first period (start of interventions), *m*_2_ is the midpoint of the second period (end of interventions) and *t* is time. Note that if *b*_1_ *> b*_2_ the function increases first and then decreases, and vice versa. To get a good set of parameters for *β*(*t*), we applied Eqn (1) to emulate infection data (from February 22 to September 1 2020) from Italy, one of the worst hit countries during the SARS-CoV-2 pandemic.

The main goal of vaccination in this model is to block transmission. Every day, there is contact between infected and vaccinated individuals. Vaccinated individuals that become exposed due to contact with an exposed individual move into the *E* state with probability (1 − *η*)*β*(*t*), where *η* is the efficacy of the vaccine. Individuals for whom the vaccine is effective remain in the *V* state whilst those for whom the vaccine is not effective move into the *E* state and later into the *I* state after which they proceed to the *R* state. Furthermore, depending on the vaccination strategy being modelled, the time of vaccination as well as population coverage varies.

##### Algorithm 1: Epidemic process on social network

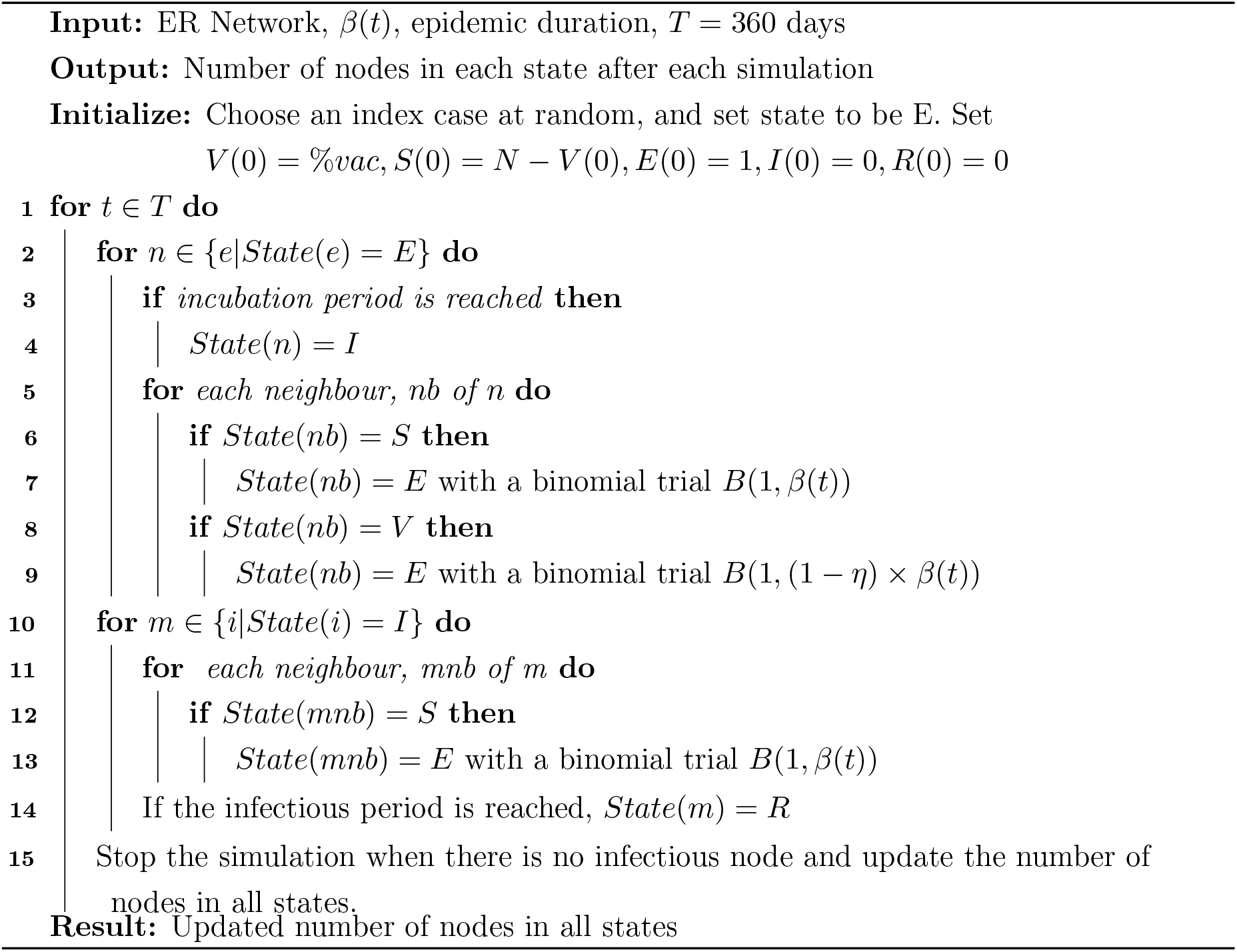

For the first strategy which we refer to as the **classical mass vaccination** strategy, a fraction of individuals start in the *V* state (denoted %*vac*) with one index case (patient zero) and the rest in the *S* state. The population coverage in this case ranges from 10%, 20%, …, 100%. The modelling process follows as in Algorithm 1.

For the second strategy which we refer to as **ring vaccination**, vaccination occurs after a percentage (1% or 3%) of the population has been exposed to the virus during the epidemic. In this case, we simulate the epidemics as described above (in Algorithm 1) with no vaccination and only begin vaccine administration after a proportion of the population (%*exposed*) has been exposed (See Algorithm 2 for more details). We assume that once a case is diagnosed, all the contacts or neighbours are traceable and vaccinated. To initiate the ring vaccination, susceptible contacts of exposed individuals can be vaccinated in a binomial trial with probability (1 − *η*)*β*(*t*). This vaccination is further extended to susceptible contacts of these first contacts with the assumption that contacts are identified through contact tracing. Contacts of contacts are also vaccinated with the same probability. In our model, we also assume that traced and identified susceptible contacts and contacts of contacts are vaccinated. This process is described in Algorithm 2 and detailed implementation can be found at Code and examples. In both strategies, we assume that the vaccine does not have an effect on infected or exposed individuals. Table 1 summarizes all parameters and key terms used in this study.

**Table 1:** Definition of key terms and parameters

| Key term | Definition | Value |
| --- | --- | --- |
| $N$ | number of nodes in the network | $10^6$ |
| $\langle k \rangle$ | average degree distribution of nodes in the network | 10 |
| transmission probability $\beta$ | the probability that infection is spread due to contact between an infected node and a susceptible node | Eqn (1) |
| incubation period | the interval between exposure to virus and initial occurrence of symptoms | 1 – 5 days |
| infection period | interval between symptom onset to recovery | 6 – 19 days |
| vaccine efficacy $\eta$ | efficacy of vaccine | varies |
| $\%vac$ | percentage of population vaccinated prior to infection (before case zero) | varies |
| $T$ | epidemic duration | 360 days |
| $\%exposed$ | percentage of population exposed to virus before onset of vaccination | varies |

### 2.2 Simulation Scenarios

Several scenarios are explored here. First, we consider an epidemic implemented with no vaccination. This scenario is evident of the epidemic outcome without the discovery of a vaccine. Subsequent scenarios consist of the overall performance of the vaccine by considering its efficacy in preventing transmission and symptomatic infection using either the classical mass vaccination or the ring vaccination strategy. In our simulations, vaccine efficacy varied between 40% − 100% whereas population coverage for classical vaccination varied between 10% − 100%. For ring vaccination, the proportion of exposed individuals (%*exposed*) varied as 1% and 3% (see Table 2). For all simulations: *N* = 10^6^, *b*_1_ = 0.028, *b*_2_ = 0.001, *r*_1_ = 0.09, *r*_2_ = 0.04, *m*_1_ = day 50, *m*_2_ = day 126 and *T* = 360 days. Each scenario is repeated 50 times and the mean of infection cases taken for analysis.

**Table 2:** Vaccination scenarios with corresponding population coverages and vaccine efficacies considered in this study.

| Scenario | Coverage | Time of vaccination | Effect of vaccine (%) |
| --- | --- | --- | --- |
| No vaccination | 0% | - | - |
| Mass vaccination | 10%, 20%, …, 100% | Before case zero | 40, 60, 80, 100 |
| Ring vaccination | 1st order and identified contacts AND contacts of contacts | After percentage of population infected (varies between 1%, 3%) | 40, 60, 80, 100 |

#### Algorithm 2 Ring vaccination model on social network

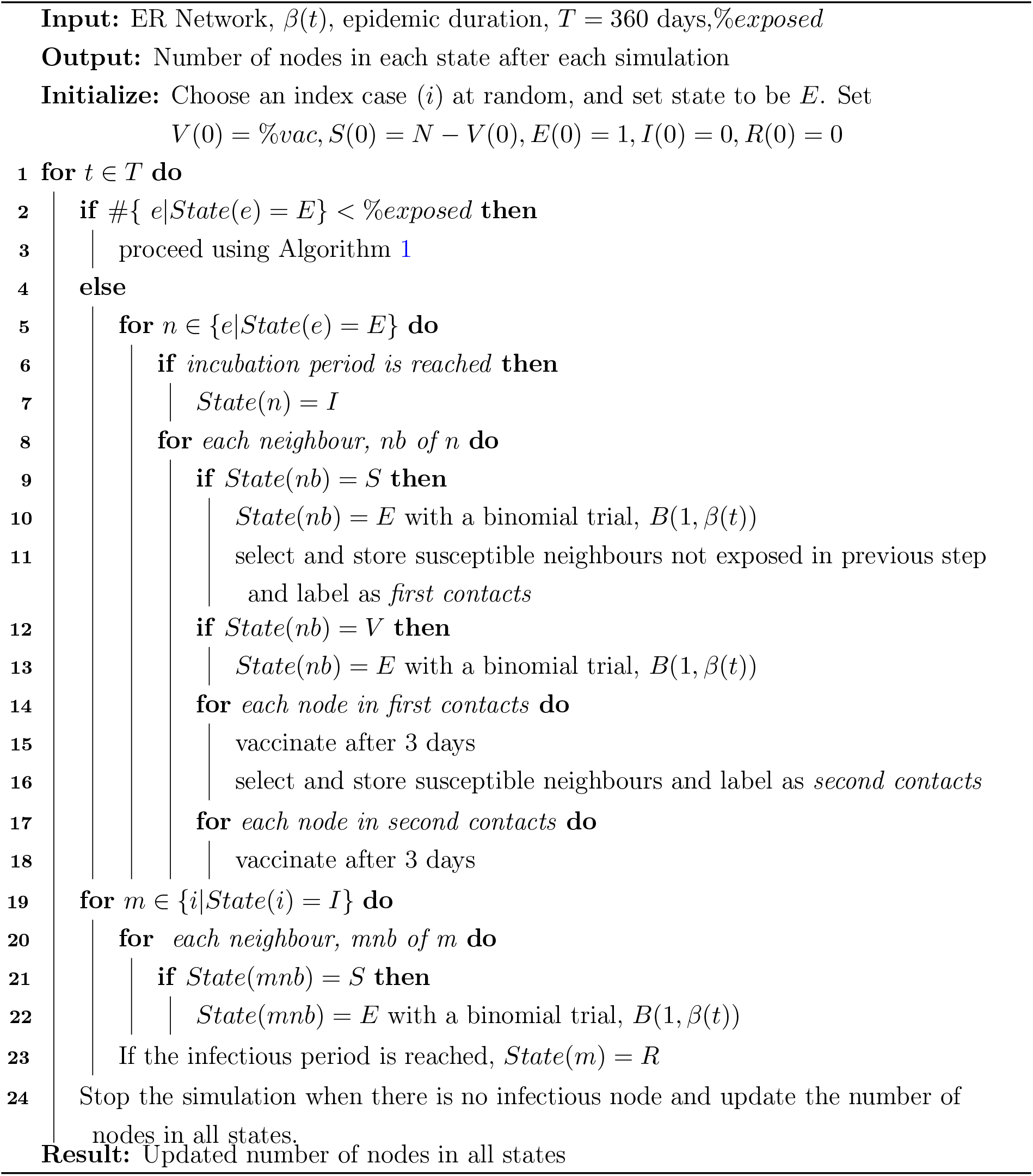

### 2.3 High-Performance Computing

Implementations of this network model was computationally demanding and challenging using conventional resources. For instance, one simulation can take up to several hours or days to complete in a modern desktop computer. Thus, due to these limitations, we employed a High-Performance Computing (HPC) cluster for our simulations. The cluster used is FUCHS-CSC from the Center for Scientific Computing (CSC, Frankfurt, Germany). It is based on 72 dual-socket AMD Magny-Cours CPU compute nodes with 64 GB of RAM, 250 dual-socket AMD Istanbul compute nodes with 32 GB of RAM and 36 quad-socket AMD Magny-Cours compute nodes with 128 GB of RAM each. A simulation in the HPC takes about 5h to complete for the classical vaccination scenarios and about 5 − 8 days for the ring vaccination scenarios. This project is coded in Python.

## 3 Results

Our parameter fitting for *β*(*t*) shows infection peaks at comparable time points with varying population percentages but essentially a qualitative fit of the data (see Figure 2). The parameter values used to fit *β*(*t*) are: *b*_1_ = 0.028, *b*_2_ = 0.001, *r*_1_ = 0.09, *r*_2_ = 0.04, *m*_1_ = 50, *m*_2_ = 126.

**Figure 2:**
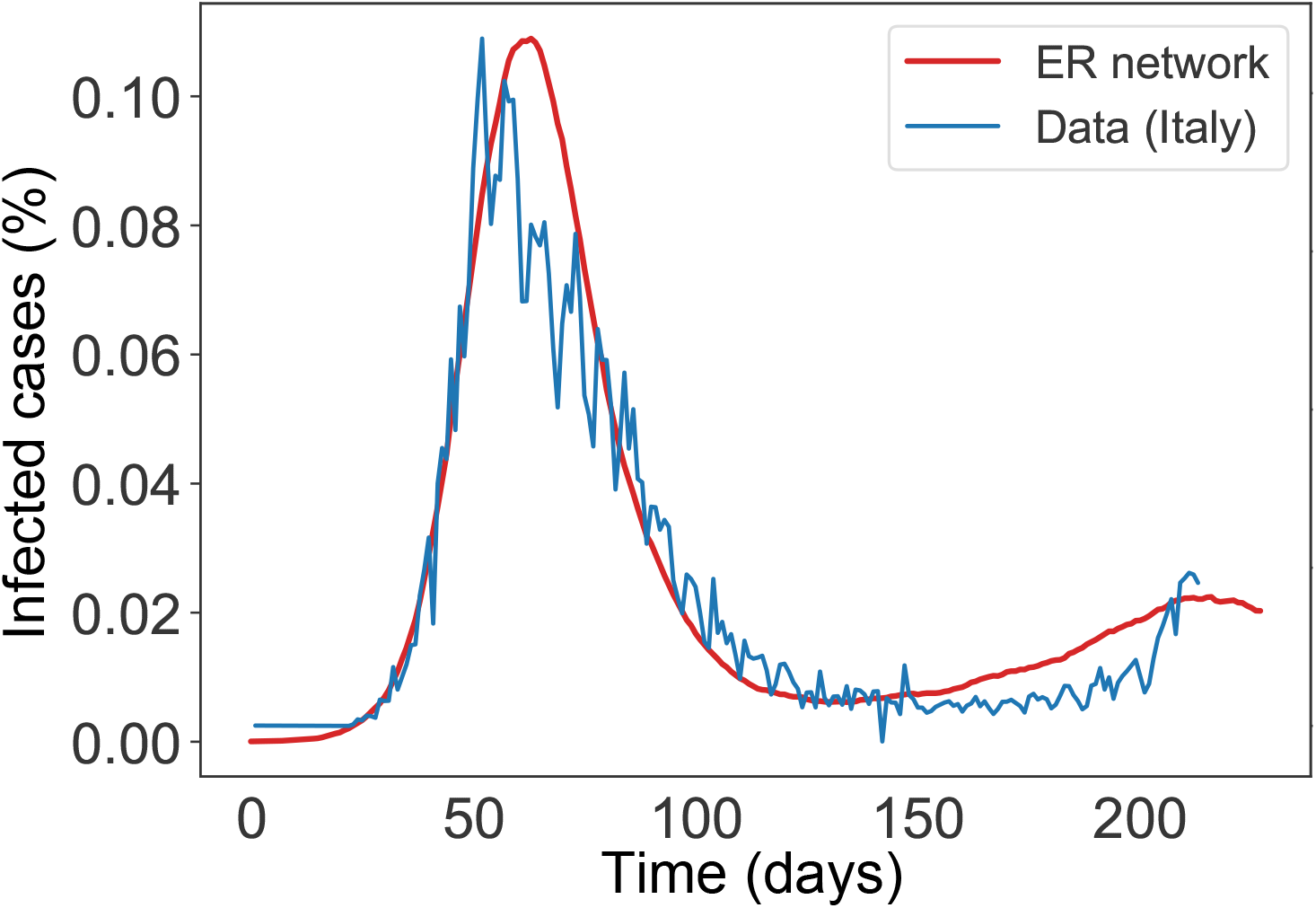
Comparism of infection cases between the network model simulation using *β*(*t*) and Italian SARS-CoV-2 data from February 22 2020 to September 1 2020. The y-axis shows the infection cases. Parameter values used are *b*_1_ = 0.028,*b*_2_ = 0.001, *r*_1_ = 0.09, *r*_2_ = 0.04, *m*_1_ = 50, *m*_2_ = 126.

### 3.1 COVID-19 dynamics - control scenario

In a completely susceptible population, the introduction of one exposed individual leads to the spread of the infection with more than one peak of cases of infection after some months. In Figure 3, we see that the mean number of infected cases first peaks in the second month after infection onset with about 12.8% of the population infected. By the fifth month during the infection course, there arises a second peak of infected cases which closely matches the first.

**Figure 3:**
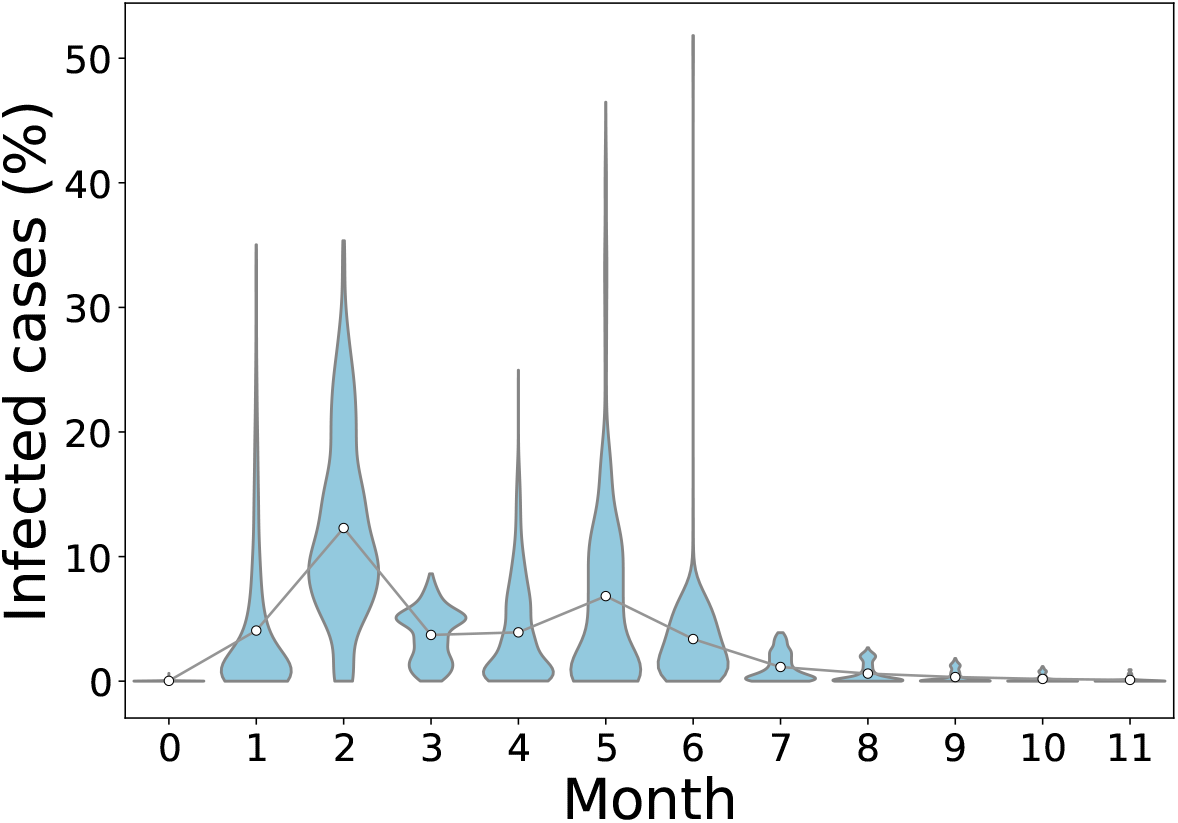
Illustration of the distribution of the final infected cases in different timing for a no vaccination scenario. A population of N= 10^6^ individuals was generated and 50 simulations were run to simulate the epidemic in the course of one year. An individual was chosen randomly as patient zero for each run. Circles represent mean infection cases for each month connected by lines.

### 3.2 Mass Vaccination Scenarios

These scenarios determine the outcome of vaccination at the initial stages of the epidemic before infected cases peak. When vaccine efficacy is 40%, a population coverage of 50% or more is needed to achieve infection peak with less than 10% infection cases. Table 3 reveals that with vaccine efficacy of 60%, a coverage more than 70% and above keeps mean infection cases on the low with elimination of the peak occurring when coverage is more than 80%. Furthermore, low cases of infection are observed when 70% or more of the population is vaccinated and vaccine efficacy is 80%. On the other hand, a vaccine which is 100% efficacious requires 60% or more of the population to be vaccinated to ensure there is no peak of infections. As seen in Figure 4 (see also Figure S1 - S4 in Supplementary Material), peak(s) of infection cases are observed when a small percentage of the population is vaccinated. For instance, in all scenarios, two infection peaks are observed when %*vac* is 40%. Similarly, there are peaks of infection for vaccine coverage between 20% and 60% even when *η* varies. Therefore, vaccinating a small proportion of the population is not useful in these instances as cases can still peak even with a vaccine with 100% efficacy. Thus, the key to eliminating infection peaks is to administer very efficacious vaccines to many individuals.

**Table 3:** Average infection peak values considering vaccine efficacy and population coverage

| No vaccination scenario<br>(control scenario) |  |  |  |  |
| --- | --- | --- | --- | --- |
| 12.79 |  |  |  |  |
| Mass vaccination scenarios<br>(vaccination before case zero) |  |  |  |  |
| Population Coverage | Vaccine Efficacy |  |  |  |
|  | 40% | 60% | 80% | 100% |
| 10% | 12.67 | 11.72 | 13.37 | 14.33 |
| 20% | 16.71 | 16.92 | 14.27 | 16.27 |
| 30% | 12.68 | 10.81 | 9.54 | 10.45 |
| 40% | 11.71 | 10.80 | 8.46 | 6.48 |
| 50% | 6.50 | 4.18 | 4.29 | 2.33 |
| 60% | 5.69 | 2.97 | 0.97 | 0.10 |
| 70% | 1.55 | $1.07e^{-3}$ | $2.78e^{-4}$ | $5.01e^{-5}$ |
| 80% | $2.15e^{-4}$ | $2.07e^{-4}$ | $1.20e^{-5}$ | $2.47e^{-5}$ |
| 90% | 0 | $4.57e^{-5}$ | $5.20e^{-6}$ | $8.67e^{-7}$ |
| 100% | 0 | 0 | 0 | 0 |
| Ring vaccination scenarios<br>(population exposed before vaccination) |  |  |  |  |
|  | Vaccine Efficacy |  |  |  |
|  | 40% | 60% | 80% | 100% |
| (1% of population exposed before vaccination) |  |  |  |  |
|  | 0.95 | 0.95 | 1.03 | 0.97 |
| (3% of population exposed before vaccination) |  |  |  |  |
|  | 2.77 | 3.45 | 3.23 | 3.06 |

**Figure 4:**
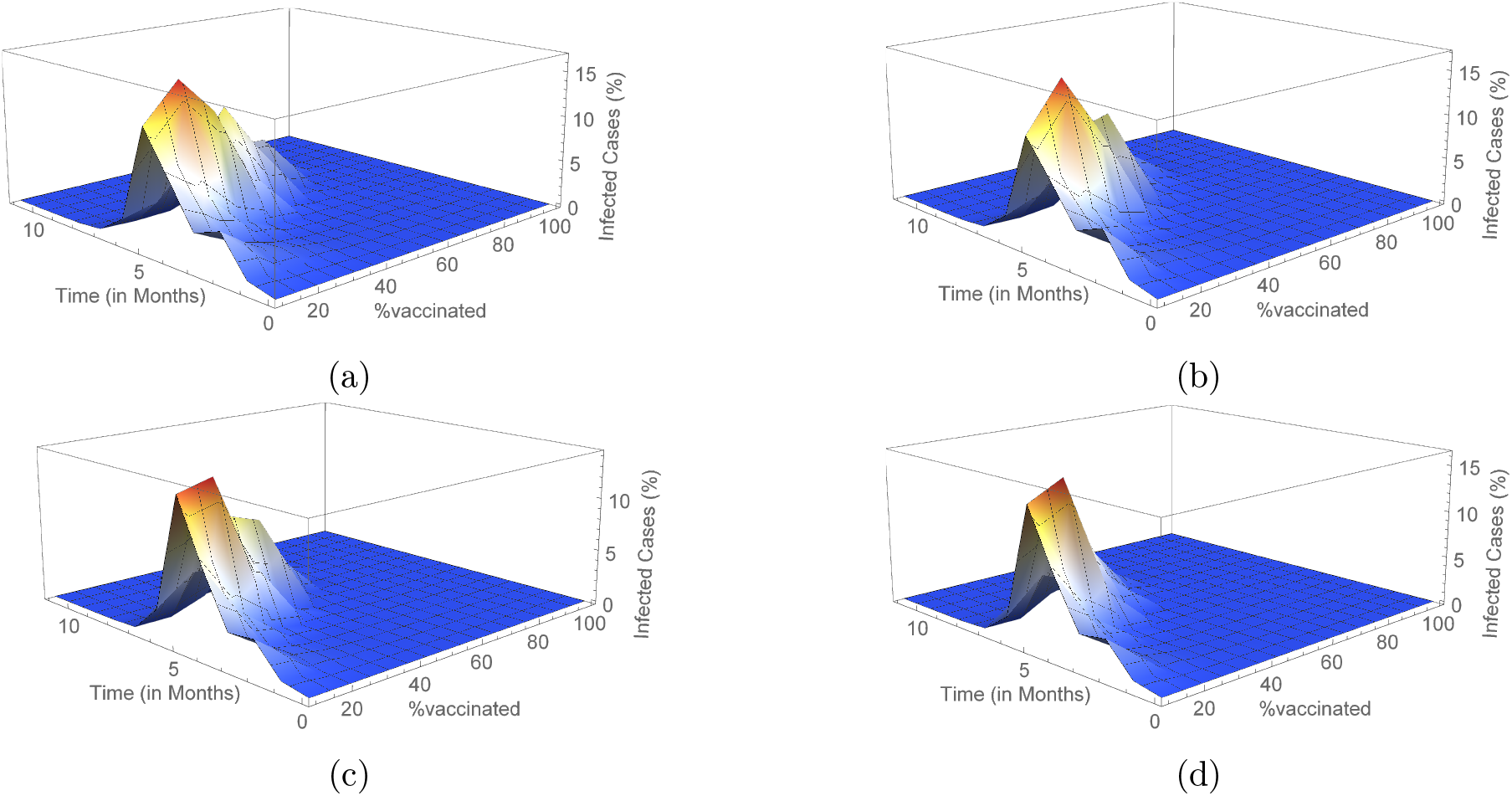
Outcome for mass vaccination scenarios for each vaccine efficacy percentage. This shows the changes in mean infected cases over time under the different vaccine efficacies. In (a), (b), (c) and (d) vaccine efficacies are 40%, 60%, 80% and 100% respectively.

### 3.3 Ring vaccination scenario

We carried out simulations using Algorithm 2 to determine the infection outcome when ring vaccination is used. In Figure 5 (see also Figure S5 and Table 4), we show these outcomes with varying scenarios of vaccine efficacy and when 1% or 3% of the population is already exposed to the disease before the onset of vaccination. We see that there are less cases realised with a 1% exposed population before the start of vaccination as compared to a 3% exposed population. In addition, the number of infected cases in these scenarios are considerably lower than that of the classical vaccination method. Also, even with a 100% efficacious vaccine, total eradication of the peak is not achieved irrespective of the exposed population prior to vaccination (See also Figure S5a and Figure S5b). Furthermore, only one peak of infection is seen using this strategy compared to the sometimes two peaks of the mass vaccination strategy.

**Table 4:** Average population coverage (in %) for ring vaccination scenarios considering vaccine efficacy.

| Vaccine Efficacy |  |  |  |
| --- | --- | --- | --- |
| 40% | 60% | 80% | 100% |
| (1% of population exposed before vaccination) |  |  |  |
| 35.94 | 36.32 | 36.30 | 36.57 |
| (3% of population exposed before vaccination) |  |  |  |
| 59.90 | 60.47 | 59.54 | 57.38 |

**Figure 5:**
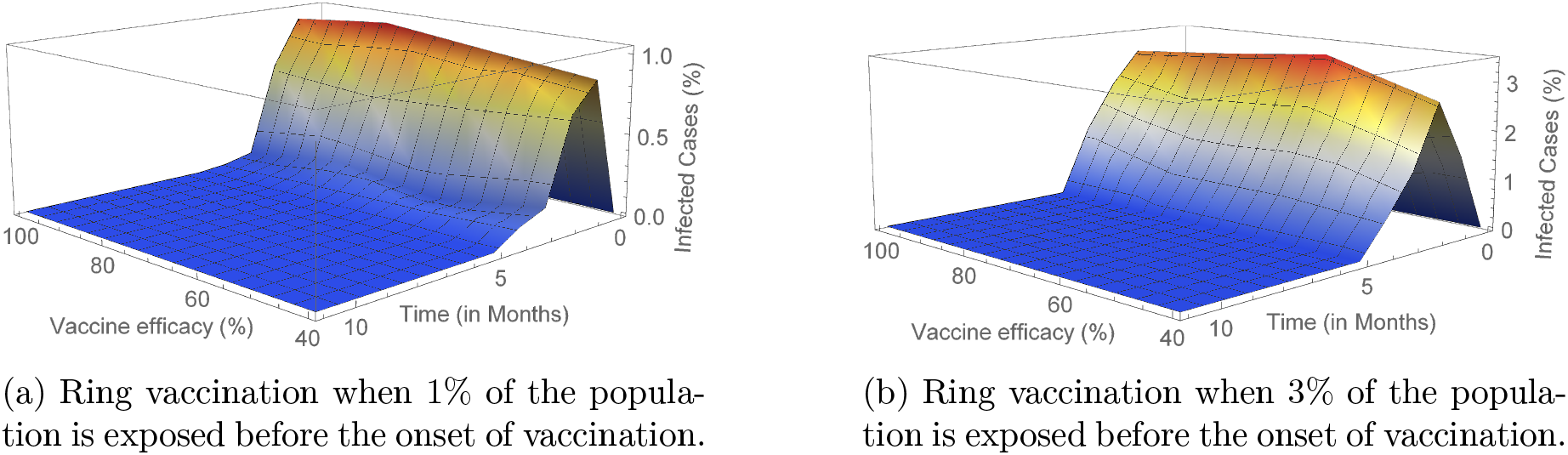
Outcome for ring vaccination when 1% and 3% of the population has been exposed before the onset of vaccination considering varying vaccine efficacy (*η* = 40%, 60%, 80%, 100%).

In comparison to mass vaccination, a lower percentage of the population has to be vaccinated when using a ring vaccination protocol in order to attain low infection cases (see Table 4). This is especially so as with effective contract tracing, more individuals can be vaccinated and thus decreasing the number of infections. It is worth noting also that even with the above results, the percentage of vaccinated individuals is lesser when only 1% of the population is exposed prior to vaccination than when the prior exposed population is 3%. Also, from Table 4 we see that the vaccinated populations are very similar in both scenarios respectively irrespective of the efficacy of the vaccine.

## 4 Discussion

With the ongoing spread of SARS-CoV-2 worldwide, pharmaceutical companies have also been in a race to produce safe and highly effective vaccines to counter transmission of the disease. As of December 2020, there were 52 vaccine candidates in clinical trials in humans, 13 of which are in Phase 3 trials (see Table 5) [60]. In November 2020, some pharmaceutical companies and institutes, including Pfizer Inc and BioNTech, Moderna, the University of Oxford (in collaboration with AstraZeneca), announced positive results from the first interim analyses of their Phase 3 vaccine trials [46, 48, 37]. Initial data released from these trials report that the vaccines manufactured by Pfizer Inc/BioNTech and Moderna both yielded 95% efficacy whereas that by University of Oxford (in collaboration with AstraZeneca) yielded 70% efficacy. On 2 December 2020, the UK medicines regulator MHRA granted a temporary regulatory approval for the Pfizer-BioNTech vaccine [43] which was under evaluation for emergency use authorization (EUA) status by the United States Food and Drug Administration [57] and approved for use on 11 December 2020 [43] in the United States.

**Table 5:**
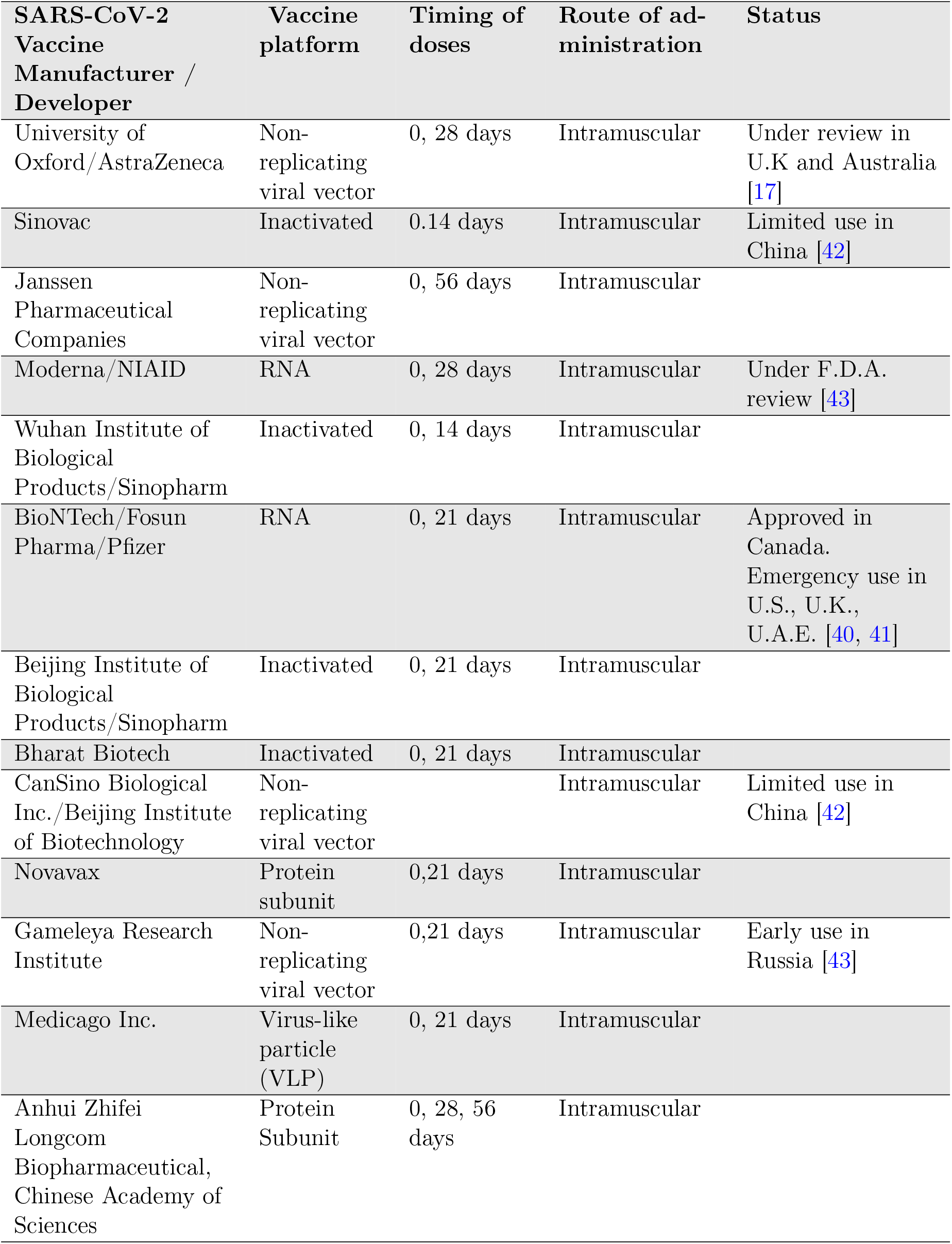
List of candidate vaccines in Phase 3 trials of clinical evaluation. Adapted from [60].

| **SARS-CoV-2**<br>**Vaccine**<br>**Manufacturer /**<br>**Developer** | **Vaccine**<br>**platform** | **Timing of**<br>**doses** | **Route of ad-**<br>**ministration** | **Status** |
| --- | --- | --- | --- | --- |
| University of<br>Oxford/AstraZeneca | Non-<br>replicating<br>viral vector | 0, 28 days | Intramuscular | Under review in<br>U.K and Australia<br>[17] |
| Sinovac | Inactivated | 0.14 days | Intramuscular | Limited use in<br>China [42] |
| Janssen<br>Pharmaceutical<br>Companies | Non-<br>replicating<br>viral vector | 0, 56 days | Intramuscular |  |
| Moderna/NIAID | RNA | 0, 28 days | Intramuscular | Under F.D.A.<br>review [43] |
| Wuhan Institute of<br>Biological<br>Products/Sinopharm | Inactivated | 0, 14 days | Intramuscular |  |
| BioNTech/Fosun<br>Pharma/Pfizer | RNA | 0, 21 days | Intramuscular | Approved in<br>Canada.<br>Emergency use in<br>U.S., U.K.,<br>U.A.E. [40, 41] |
| Beijing Institute of<br>Biological<br>Products/Sinopharm | Inactivated | 0, 21 days | Intramuscular |  |
| Bharat Biotech | Inactivated | 0, 21 days | Intramuscular |  |
| CanSino Biological<br>Inc./Beijing Institute<br>of Biotechnology | Non-<br>replicating<br>viral vector |  | Intramuscular | Limited use in<br>China [42] |
| Novavax | Protein<br>subunit | 0,21 days | Intramuscular |  |
| Gameleya Research<br>Institute | Non-<br>replicating<br>viral vector | 0,21 days | Intramuscular | Early use in<br>Russia [43] |
| Medicago Inc. | Virus-like<br>particle<br>(VLP) | 0, 21 days | Intramuscular |  |
| Anhui Zhifei<br>Longcom<br>Biopharmaceutical,<br>Chinese Academy of<br>Sciences | Protein<br>Subunit | 0, 28, 56<br>days | Intramuscular |  |

Although the Pfizer, Moderna and University of Oxford/AstraZeneca vaccines are expected to be used worldwide in coming months, in other countries such as China and Russia, other vaccines (from Sinovac in China and Gameleya Research Institute in Russia) have been developed and are currently in limited use in these countries. Table 5 summarizes the candidate vaccines under phase 3 trial and their current usage status as well as their manufacturers and vaccine platform (that is, a system which uses certain basic components as the backbone but is relatively flexible and can be adapted quickly to be used against different pathogens [24]).

In this study, we modelled the spread of SARS-CoV-2 infection using an SEIRV model structure on a social network. We run stochastic simulations to determine vaccine efficacy and population coverage limits capable of extinguishing the disease. We considered two vaccination strategies, each with varying scenarios regarding vaccine efficacy and population coverage. We found that, the introduction of a single infected person into a completely susceptible population leads to the spread of infection giving rise to more infected cases and subsequently more than one infection peak. This is an indication that in the absence of a good enough vaccine, there is a much higher chance of more than one infection peak which is already being observed in some countries [8, 6].

In addition, the introduction of vaccination lowered the number of infected cases irrespective of the type of vaccination. Our simulations and analysis show that when using a mass vaccination strategy, the lower the efficacy of the vaccine, the more people needed to be vaccinated against the disease in order to eliminate infection peak(s). For instance, a vaccine with an efficacy of 40% will require more than 80% of the entire population to be vaccinated in order to reach low infection peak values whereas a 100% effective vaccine will require more than 60% of the population to be vaccinated. Given the experimental nature and limited initial supply of vaccines, a classical mass vaccination campaign might not be feasible in many countries. However, ring vaccination of likely case contacts and contacts of cases could provide an effective alternative in distributing the vaccine to ensure low levels of infection and subsequently preventing infection peaks. This is with the assumption of effective contract tracing of infected people.

Researchers, policy makers and the general public are of the opinion that the discovery of a vaccine will allow the return to normality of life before the SARS-CoV-2 pandemic. It is worth noting that, the discovery of a proven-to-be effective vaccine however, might not reduce transmission completely. This is due to the fact that a vaccine which effectively reduces the severity of transmission does not necessarily reduce virus transmission to a comparable degree.

It is also important to consider the potential impact of voluntary mass vaccination in the efforts of clearing the epidemic [45, 49, 31, 11]. In years past, the roll-out of vaccination has been faced with declining vaccine confidence in the general public which could possibly lead to hesitancy in getting vaccinated [12]. Such instances could easily lead to a disruption of people receiving a vaccine voluntarily thus, having a detrimental effect on efforts to eradicate the disease and hence such situations should not be underestimated.

## Limitations and future work

Though exploring the effects of SARS-CoV-2 transmission on social networks, in this study we have limited our analysis to one type of social network which is the Erdos-Renyi network. To proceed towards increasing practicality, the analysis performed in this study can be extended to consider disease spread and vaccination on other social networks, such as the Barabási–Albert network [5] and small-world network [58] models, using similar scenarios and protocols. Such network models can be compared with each other and the outcomes analysed. Data on known social networks such as contact matrices and mixing patterns can also be used for further analysis and evaluation.

Simulations in this study were performed with the assumption of an optimistic condition in which there was vaccination of all contacts and contacts of contacts. In reality, contact tracing is especially difficult in the course of an ongoing epidemic and thus reduces the impact of vaccination efforts [34, 29]. Also, the model used here assumes there is equal mixing of individuals and their neighbours in the network whereas in reality this is not the case and consequently infection cases could be lower and population coverage may be reduced.

The model also assumes that all infected people recover from the disease and are immune. The effect of mortality on the dynamics of the disease is not considered as this model aims to study the general transmission dynamics and the effects of varying vaccination strategies. Within-host dynamics such as the viral dynamics and immune responses would be important to be included in a framework with higher resolution [44].

Epidemiological models of disease spread and transmission generally take into account the reproduction number (*R*_0_) of the disease to gain knowledge on the transmission process. Estimates of *R*_0_ vary widely as data continue to emerge in an ongoing epidemic. This study however, does not examine this parameter since it does not directly impact the transmission dynamics.

In summary, this study describes the transmission dynamics of SARS-CoV-2 on a social contact network and the vaccine efficacy thresholds needed to prevent new waves of the disease. For future work, we will focus on other network models and their analysis with respect to SARS-CoV-2. We will also consider the transmission potential based on *R*_0_ and the dynamics in this regard.

## Data Availability

The code and the data-sets supporting this article have been uploaded as electronic supplementary material.

https://github.com/Josephine-Tetteh/COVID-19-Network-Model.git

## Supplementary materials

**Figure S1:**
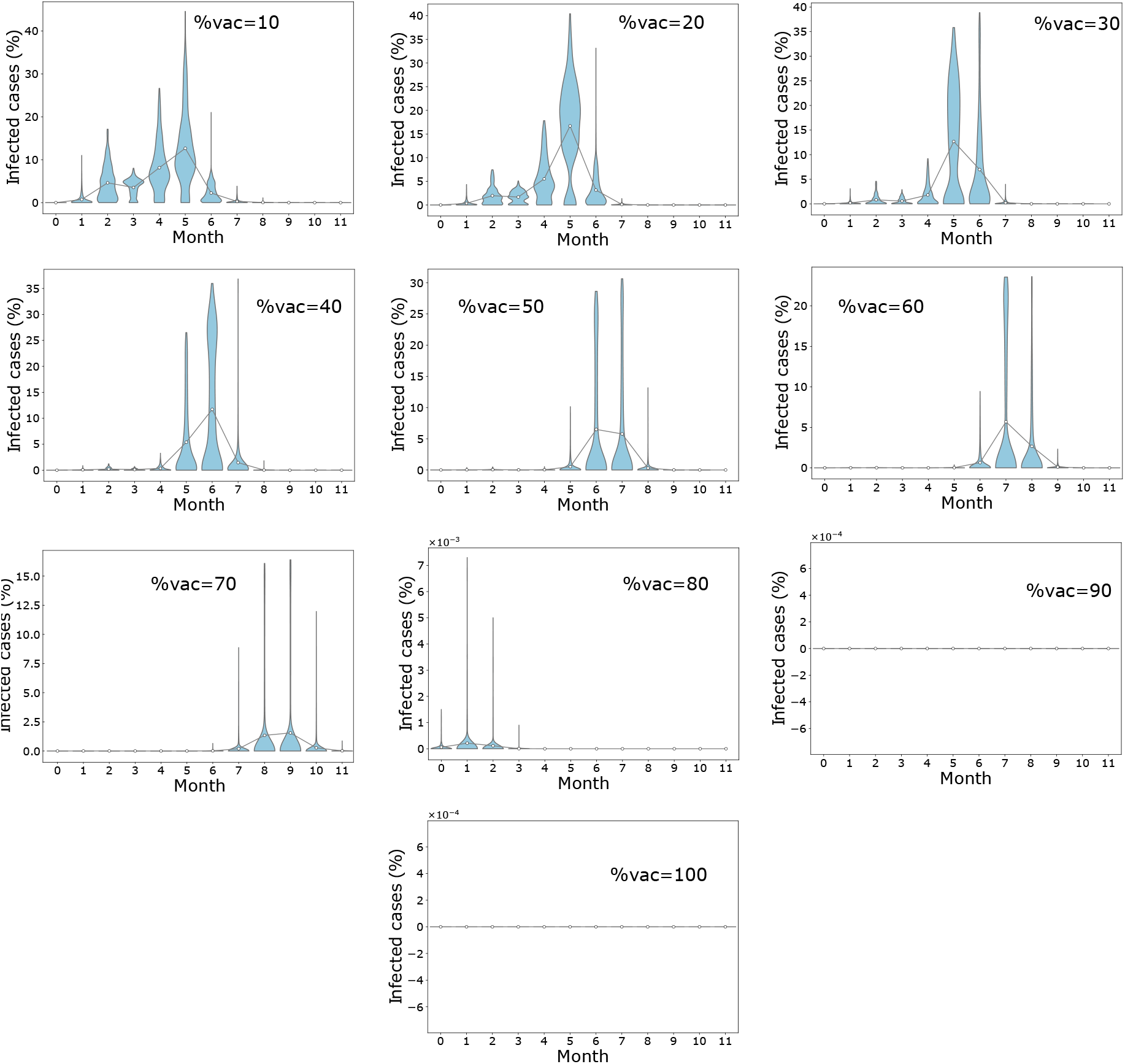
Distribution of final infected cases in different timing for mass vaccination scenario when *η* = 40%. Population coverage varies from 10% 100%. Circles represent mean infection cases for each month connected by lines.

**Figure S2:**
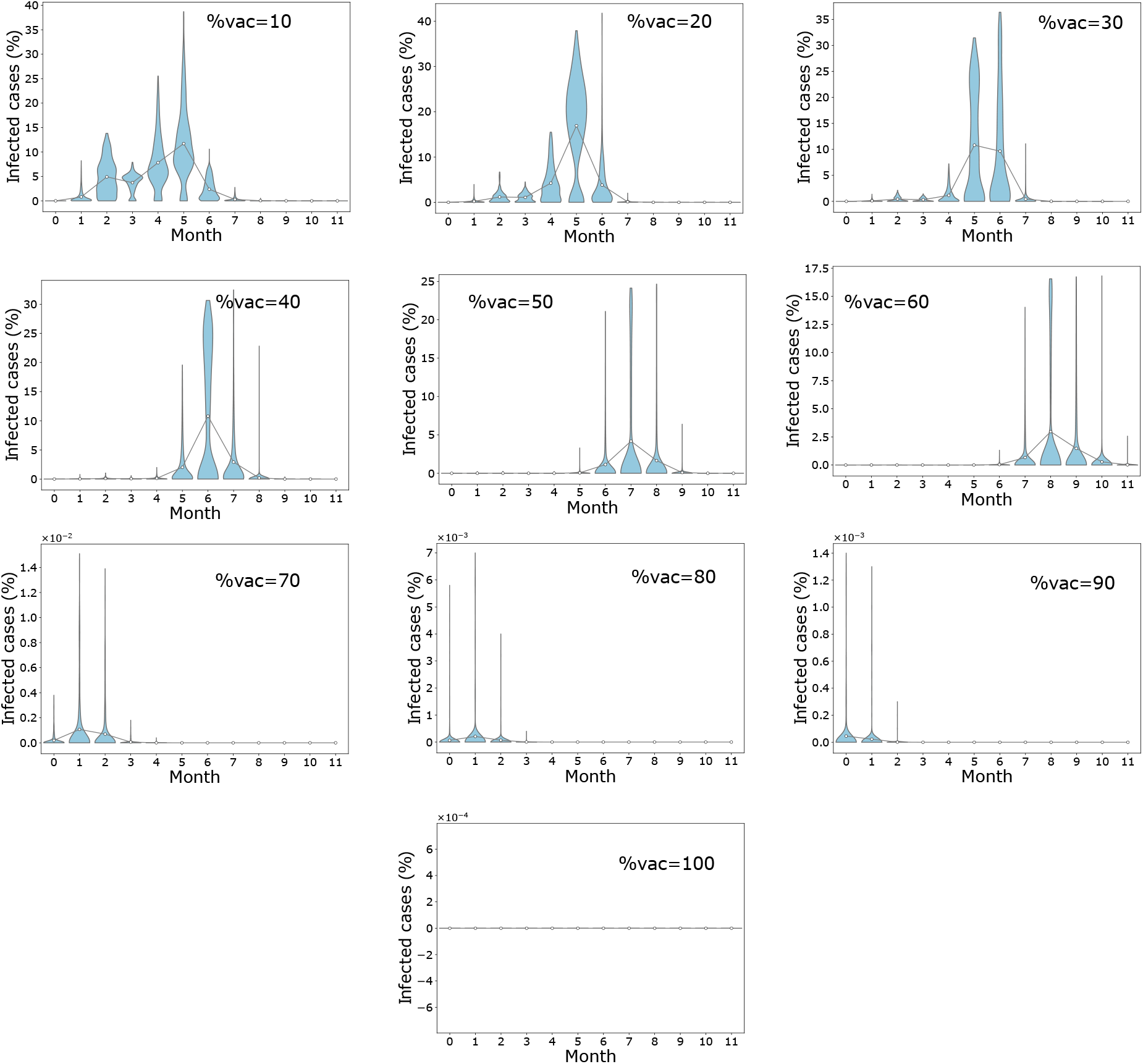
Distribution of final infected cases in different timing for mass vaccination scenario when *η* = 60%. Population coverage varies from 10% 100%. Circles represent mean infection cases for each month connected by lines.

**Figure S3:**
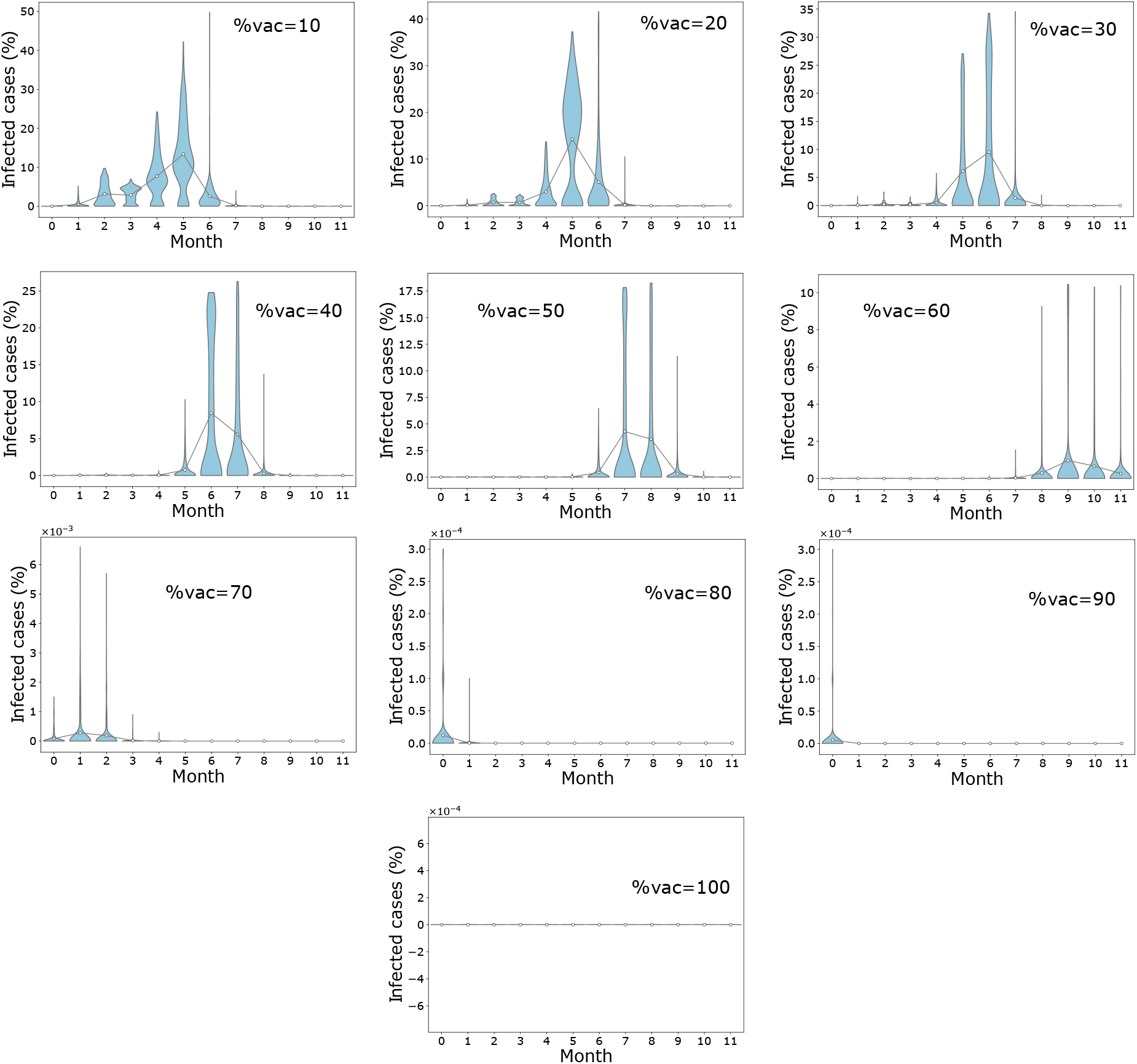
Distribution of final infected cases in different timing for mass vaccination scenario when *η* = 80%. Population coverage varies from 10% 100%. Circles represent mean infection cases for each month connected by lines.

**Figure S4:**
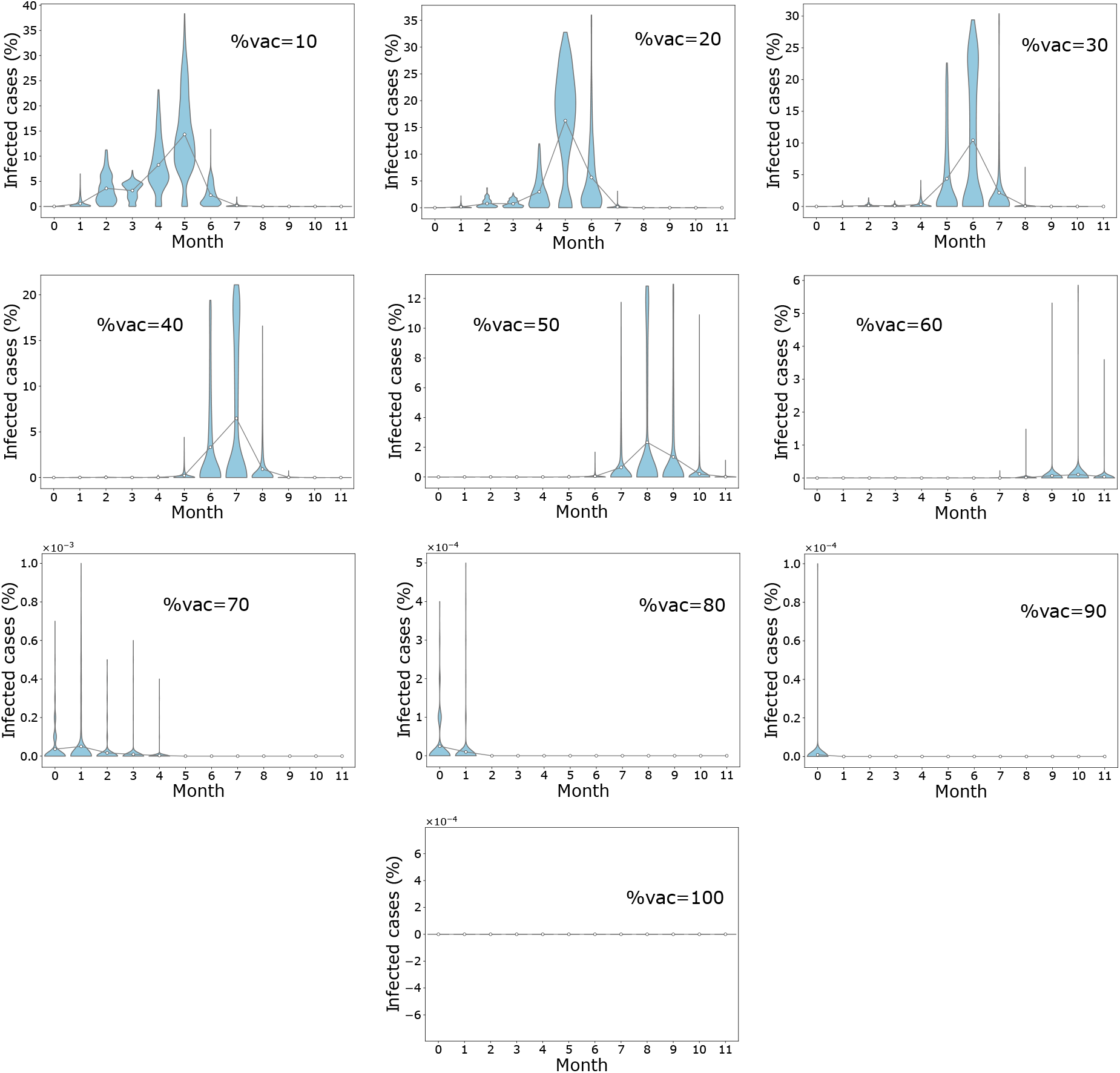
Distribution of final infected cases in different timing for mass vaccination scenario when *η* = 100%. Population coverage varies from 10% 100%. Circles represent mean infection cases for each month connected by lines.

**Figure S5:**
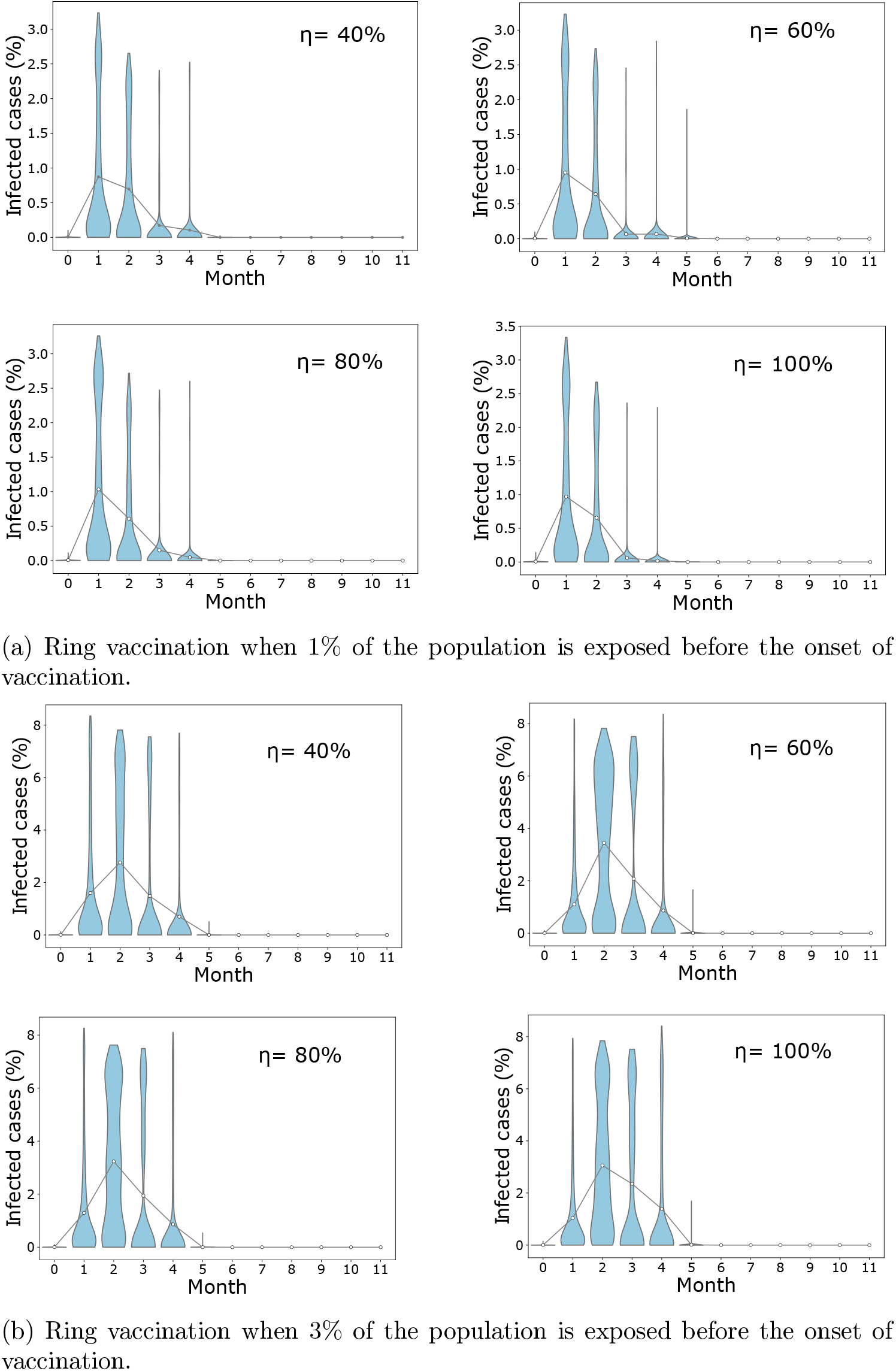
Illustrating number of infected cases when ring vaccination is employed and when 1% and 3% of the population has been exposed before the onset of vaccination considering varying vaccine efficacies (*η* = 40%, 60%, 80%, 100%).

## Notes

### Competing Interest Statement

The authors have declared no competing interest.

### Funding Statement

This work is supported by the Deutsche Forschungsgemeinschaft (HE7707/5-1),
the Alfons und Gertrud Kassel-Stiftung and the Mexican National Science and Technology Council (CONACYT)

